# Photograph-Based Skeletal Malocclusion Screening with Geometric Machine Learning: External Validation Across Turkish and Non-European Cohorts

**DOI:** 10.64898/2026.10.28.26366413

**Authors:** Aslı Eker Davut, Mehmet Cihan Sakman, Ahmet Yağcı, Berrak Çakmak, Emmanuel Jr Dumu, Yunus Emre Uçaker, Cenk Ahmet Akcan, Özgür Kılıç, Tuğba Önal-Süzek, Banu Kılıç

**Affiliations:** Department of Orthodontics, Institute of Health Sciences, Bezmialem Vakif University, Istanbul, Turkey; Institute of Data Analysis and Process Design, Zurich University of Applied Sciences (ZHAW), Winterthur, Switzerland; Department of Orthodontics, Erciyes University, Kayseri, Turkey; Department of Orthodontics, Gazi University, Ankara, Turkey; PEBS Orthodontics, Brussels, Belgium; Department of Orthodontics, Hacettepe University, Ankara, Turkey; Department of Computer Engineering, Muğla Sıtkı Koçman University, Muğla, Turkey; Kedi Mobil Uygulama Anonim Şirketi, Istanbul, Turkey; Department of Orthodontics, Bezmialem Vakif University, Istanbul, Turkey

**Keywords:** calibration, external validation, generalizability, machine learning, orthodontic screening, skeletal malocclusion

## Abstract

A photograph-based screening classifier for skeletal malocclusion maps 100 engineered geometric feature vectors, derived from a side-profile smartphone photograph, to orthodontic class labels, enabling non-invasive triage without radiographic imaging. This study reports external validation across two independent cohorts that contributed no cases to training. Two binary tasks were evaluated; Class III against non-Class III, and Class I against Class II; each with its own development set and model, applied without modification at the default threshold. One external cohort was Turkish, drawn from an institution not represented in the development set; the other was a non-European cohort from a Belgian private orthodontic clinic. Sensitivity, specificity and the area under the receiver operating characteristic curve (AUC) were pre-specified as primary outcome measures, since none depends on outcome prevalence. In the Class III task, sensitivity was similar across all three evaluation sets (0.801, 0.836, 0.848; all adjusted p>0.7), while specificity fell from 0.766 in the Turkish external cohort to 0.697 in the non-European cohort. AUC was 0.902 and 0.835 respectively (difference 0.066; p=0.029). In the Class I and Class II task the corresponding values were 0.875 and 0.723 (difference 0.152; p<0.001). A subgroup analysis showed lower sensitivity in children than adults across both cohorts (0.71–0.80 versus 0.89–0.90). AUC was lower in the non-European cohort for all three models in both tasks. The observed performance differences are consistent with a population-specific generalizability effect rather than model-specific failure and support mandatory external validation on target populations before clinical deployment.

## I. Introduction

ARTIFICIAL intelligence has come into widespread use in medicine [1], [2], in dentistry [3], and in orthodontics for diagnosis, treatment planning and patient monitoring [4], [5]. Cephalometric analyses used in routine practice can now be performed automatically. Artificial intelligence models are also available that classify patients according to skeletal and dental relationships [4]. One such system estimates the skeletal relationship from a side-profile photograph taken on a standard smartphone, without requiring radiographic imaging or specialist equipment [6]. The machine learning models evaluated in this study are deployed in two free-to-use interfaces operated by Kedi Mobil Uygulama Anonim Şirketi: a mobile application available on iOS, Android and Samsung Galaxy Store, and a web-based screening interface [6]. Both are provided free of charge as public awareness tools and generate no revenue from the screening function of the machine learning model.

Early orthodontic diagnosis matters clinically. In skeletal Class III malocclusion, orthopedic correction decision of the orthodontist depends on the orthodontist’s prediction of the remaining future craniofacial growth [7], [8]. Class II malocclusion is likewise addressed by growth modification, which depends on remaining growth potential [9]. The American Association of Orthodontists recommends that children be screened by an orthodontist at age seven [10].

The consequences of skeletal discrepancy are not confined to childhood. A meta-analysis of cephalometric studies in adults with obstructive sleep apnea found a more retrusive and shorter mandible than in controls, together with reduced pharyngeal airway space [11]. Adults with skeletal Class III malocclusion report poorer oral health-related quality of life than those with Class I, across physical, psychological and social domains [12]. Preliminary assessment may therefore have value beyond the growth period. Access to an orthodontic examination remains limited, particularly in rural and underserved regions of the world. A system of this kind may therefore help direct patients towards examination where access is limited.

Assessment from a photograph rests on the relationship between facial morphology and the underlying skeleton. This relationship may not be identical across populations. Turkish adults differ from European-American adults in facial convexity, upper and lower lip position, chin prominence and chin thickness [13]. Turkish adolescents likewise show smaller midfacial and mandibular lengths and more retrusive faces than their North American counterparts [14]. A relationship learned in one population may therefore not hold in another, even when the skeletal criterion is unchanged. Such a difference would be expected to produce a particular pattern of failure outside the development population. Sensitivity would be largely preserved, because the features associated with the target class remain present. Specificity would decline, because some unaffected individuals would fall within the range of measurements the model has learned to associate with the target class.

External validation is uncommon in orthodontic artificial intelligence studies, and validation across populations is rarer still [15]. In medical AI, cross-population performance differences are often attributed to the training data composition of the machine learning model [16]. A difference in outcome prevalence between development and validation settings can also alter predictive values without any change in model behavior. Separating these requires prevalence to be controlled by design and prevalence independent measures to be reported. This study evaluated the generalizability of a machine learning based orthodontic diagnostic model across two external cohorts, one drawn from the development population and one from a different population, using prevalence independent outcome measures.

## II. Materials and methods

This retrospective diagnostic accuracy study was approved by the Non-Interventional Research Ethics Committee of Bezmialem Vakıf University (decision numbers 2024/342 on 04 September 2024 and 2024/438 on 11 December 2024). This study is registered on ClinicalTrials.gov (Identifier: NCT07162753, registered on 26 August 2025). It was conducted in accordance with the Declaration of Helsinki, and all data were anonymized. All data were retrospective and anonymized, and their use was covered by the ethics approvals cited above. Patients at the Belgian clinic had consented to the use of their visual data in scientific studies.

The model was evaluated in two binary classification tasks. In the first, skeletal Class III was distinguished from non-Class III, with Class I and Class II combined into a single negative category. In the second, Class I was distinguished from Class II. Including a second task with a different class structure, a different development dataset, and a different top ranked model provides a within study replication, since a cross-population pattern specific to one task or one classifier would not be expected to recur. If it appears in both, it is more likely to reflect a property of the approach rather than an artefact of a single experimental setting. Each task had its own development set, feature selection and model ranking.

Skeletal classification was established from pre-treatment lateral cephalograms using the ANB angle. Values of ANB < 0° were classified as Class III, 0° ≤ ANB ≤ 4° as Class I, and ANB > 4° as Class II. All cephalograms were traced by orthodontists, and the same thresholds and reading protocol were applied in every cohort. In the Belgian non-European external cohort, the readings were performed by two orthodontists. The ANB angle was used only to derive the reference standard and was never available to the model as an input.

Photographs were collected at three university orthodontic departments in Turkey from patients aged 8 to 35 years. The three departments contributed 796, 1,489 and 681 cases to the Class III development set and 1,472, 2,761 and 1,359 to the Class I and Class II development set. Inclusion required a lateral profile photograph taken in a standardized natural head position against a plain background before orthodontic treatment began, together with a pre-treatment lateral cephalogram. Patients with congenital craniofacial deformity, facial swelling, previous orthodontic treatment, or difficulty following positioning instructions were excluded [6].

Before balancing, duplicate records were identified by matching feature vectors to six decimal places and removed. Records appearing under conflicting class labels were also discarded, because their correct label could not be determined. This left 7,885 of 7,924 records for the Class III task (39 duplicates, 11 of them cross-class) and 6,410 of 6,442 for the Class I and Class II task (32 duplicates, 8 of them cross-class). Development sets of 2,966 cases (1,478 Class III, 1,488 non-Class III) and 5,592 cases (2,796 per class) were drawn from these pools. In the Class III task the negative group was drawn in equal numbers from Class I and Class II. The larger class was down sampled to match the smaller class in size and in age distribution simultaneously. Adults comprised 60% of both classes in both tasks, with a residual between-class difference of 0.0%. Each development set was divided in an 80:20 ratio into training and internal validation partitions. This division preceded feature selection.

The Turkish external cohort comprised patients attending an independent Turkish university orthodontic department not represented in the development set. It contained 256 cases for the Class III task, balanced by class, and 190 for the Class I and Class II task. The Belgian non-European external cohort was recruited at a private orthodontic clinic in Brussels. For the Class III task, 274 cases were drawn from a source cohort of 873 patients using a fixed random seed (seed = 42). These comprised 132 Class III cases and 142 non-Class III cases, the latter drawn in equal numbers from Class I and Class II. Class III prevalence was therefore 48.2%, against 49.8% in internal validation, and age was matched between classes with a residual difference of 0.3%. For the Class I and Class II task the full available cohort of 706 cases was used. Neither external cohort contributed any case to model training. Both were evaluated using the same saved model files, at the unchanged default decision threshold. Cohort characteristics are summarized in Table I.

**TABLE I.** Cohort characteristics.

|  | Class III task |  |  | Class I and II task |  |  |
| --- | --- | --- | --- | --- | --- | --- |
|  | Development | Turkish ext. | Belgian non-European | Development | Turkish ext. | Belgian non-European |
| Source pool, n | 7,924 | 320 | 873 | 6,442 | 190 | 706 |
| Analysis set, n | 2,966 | 256 | 274 | 5,592 | 190 | 706 |
| Internal validation, n | 594 |  |  | 1,119 |  |  |
| Positive class, n (%) | 1,478 (49.8) | 128 (50.0) | 132 (48.2) | 2,796 (50.0) | 136 (71.6) | 390 (55.2) |
| Balanced by class | Yes | Yes | Yes | Yes | No | No |
| Age matched | Yes (0.0%) | No (18.8%) | Yes (0.3%) | Yes (0.0%) | n/r | n/r |
| Adults, % | 60 | 58 | 46 | 60 | n/r | n/r |
| Bezmialem, n | 796 | — | — | 1,472 | — | — |
| Erciyes, n | 1,489 | — | — | 2,761 | — | — |
| Gazi, n | 681 | — | — | 1,359 | — | — |
| Contributing sites | 3 | 1 | 1 | 3 | 1 | 1 |

The positive class is Class III in the Class III task and Class II in the Class I and Class II task. Age matching refers to the residual difference in the proportion of adults between classes. Site counts are for the balanced development set; Class III prevalence at the three sites was 51.5%, 51.2% and 44.8%. n/r, not recorded.

Base facial measurements were extracted automatically from each photograph as normalized ratios of facial areas and angles, chosen for their independence from camera distance and environmental conditions, and normalized against ten reference distances defined by an orthodontist [6]. Pairwise ratios and products of the base measurements generated 13,861 candidate features. The 300 most strongly correlated with the target were retained, those correlating above 0.95 with an already-selected feature were dropped, and 100 were carried forward. Feature selection used training rows only. A convolutional neural network with a VGGFace and VGG16 backbone, trained by transfer learning and fine tuning on the same development data, was evaluated as an end to end alternative on the internal validation partition and underperformed the geometric feature approach. This outcome is consistent with reports that classical classifiers trained on domain engineered tabular features match or exceed deep learning at training set sizes of the order available here [17], [18]. Geometric features encode knowledge about the relationship between facial landmarks and the underlying skeleton as explicit prior information; end to end learning must recover this relationship from pixel level data, which requires substantially larger training corpora to generalize comparably [19].

The comparison included fourteen classification algorithms, among them a dummy model indicating chance level. Algorithms were compared by tenfold cross validation and ranked by AUC. Linear discriminant analysis ranked first in the Class III task and logistic regression in the Class I and Class II task. The three highest ranked models in each task were retained and applied unchanged to both external cohorts.

### A. Statistical Analysis

Sensitivity (the proportion of affected patients correctly identified) and specificity (the proportion of unaffected patients correctly identified) are each calculated within a single true class, so neither depends on how common the condition is in the cohort. The AUC, the probability that a randomly selected affected patient receives a higher score than a randomly selected unaffected one, depends on neither the decision threshold nor prevalence. Because none of these three measures depends on prevalence, they were pre-specified as the primary outcomes. Accuracy, precision, the F1 score and the Matthews correlation coefficient (MCC), which do depend on prevalence, were reported as secondary measures. The F1 score additionally ignores specificity, since it combines only sensitivity and precision.

Two-sided 95% confidence intervals for proportions were calculated by the Wilson score method [20]. Differences in sensitivity and specificity between cohorts were expressed as absolute differences, with confidence intervals by the Newcombe hybrid score method [21], and were tested by two proportion z tests. Bonferroni correction was applied for the three pairwise comparisons made for each measure.

Differences in AUC between the two external cohorts were estimated by bootstrap resampling with 10,000 replicates and a fixed seed, with percentile confidence intervals and two-sided p values.

Calibration was assessed by the Expected Calibration Error (ECE) and the Maximum Calibration Error (MCE) [22]. Predictions were partitioned into probability bins. Balanced accuracy was evaluated across the full range of decision thresholds on the internal validation partition of each task. Analyses were performed in Python 3.10 using scikit-learn [23], PyCaret, scipy.stats and statsmodels.

## III. Results

In the Class III task, linear discriminant analysis (LDA) ranked first of fourteen algorithms by cross validated AUC at 0.840, followed by logistic regression (LR) at 0.827 and gradient boosting (GB) at 0.820. The full ranking is given in Table II. The dummy model reached 0.500. On the internal validation partition, LDA achieved an accuracy of 0.774 against a majority-class rate of 50.2%, an AUC of 0.845, and an ECE of 0.039.

**TABLE II.** Algorithm ranking by ten fold cross validated auc.

| Rank | Model | Class III task | Class I and II task |
| --- | --- | --- | --- |
| 1 | Linear discriminant analysis | 0.840 | 0.724 |
| 2 | Logistic regression | 0.827 | 0.729 |
| 3 | Gradient boosting | 0.820 | 0.714 |
| 4 | Random forest | 0.816 | 0.705 |
| 5 | Extra trees | 0.812 | 0.703 |
| 6 | Light gradient boosting | 0.812 | 0.696 |
| 7 | Ada boost | 0.805 | 0.704 |
| 8 | Quadratic discriminant analysis | 0.794 | 0.709 |
| 9 | Naive Bayes | 0.776 | 0.704 |
| 10 | K nearest neighbours | 0.751 | 0.660 |
| 11 | Decision tree | 0.646 | 0.575 |
| 12 | Dummy model | 0.500 | 0.500 |

Algorithms are ranked by Class III task AUC. In the Class I and Class II task, logistic regression ranked first with AUC 0.729. Ten fold cross validation on the training partition of each task. Ridge and support vector models do not produce probability estimates and are omitted from the ranking.

In the Class I and Class II task, LR ranked first with a cross validated AUC of 0.729. On the internal validation partition, it achieved an accuracy of 0.655 against a majority-class rate of 50.0%, an AUC of 0.706, and an ECE of 0.038. Discrimination was lower than in the Class III task throughout.

Balanced accuracy reached its maximum at the default cut-off of 0.5 in the Class III task and in the Class I and Class II task. Class III task results are given in Table III. Sensitivity was 0.801 (95% CI 0.751 to 0.842) on internal validation, 0.836 (0.762 to 0.890) in the Turkish external cohort and 0.848 (0.778 to 0.900) in the Belgian non-European external cohort. No pairwise difference approached significance (all adjusted p ≥ 0.72).

**TABLE III.**
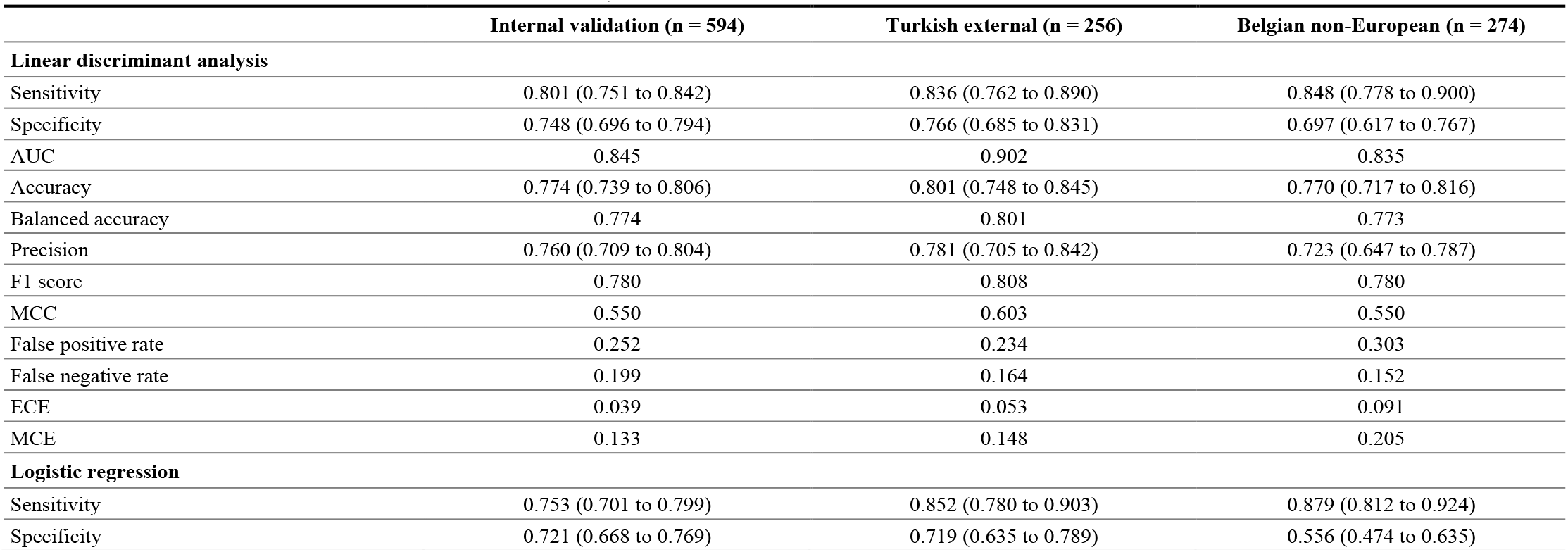
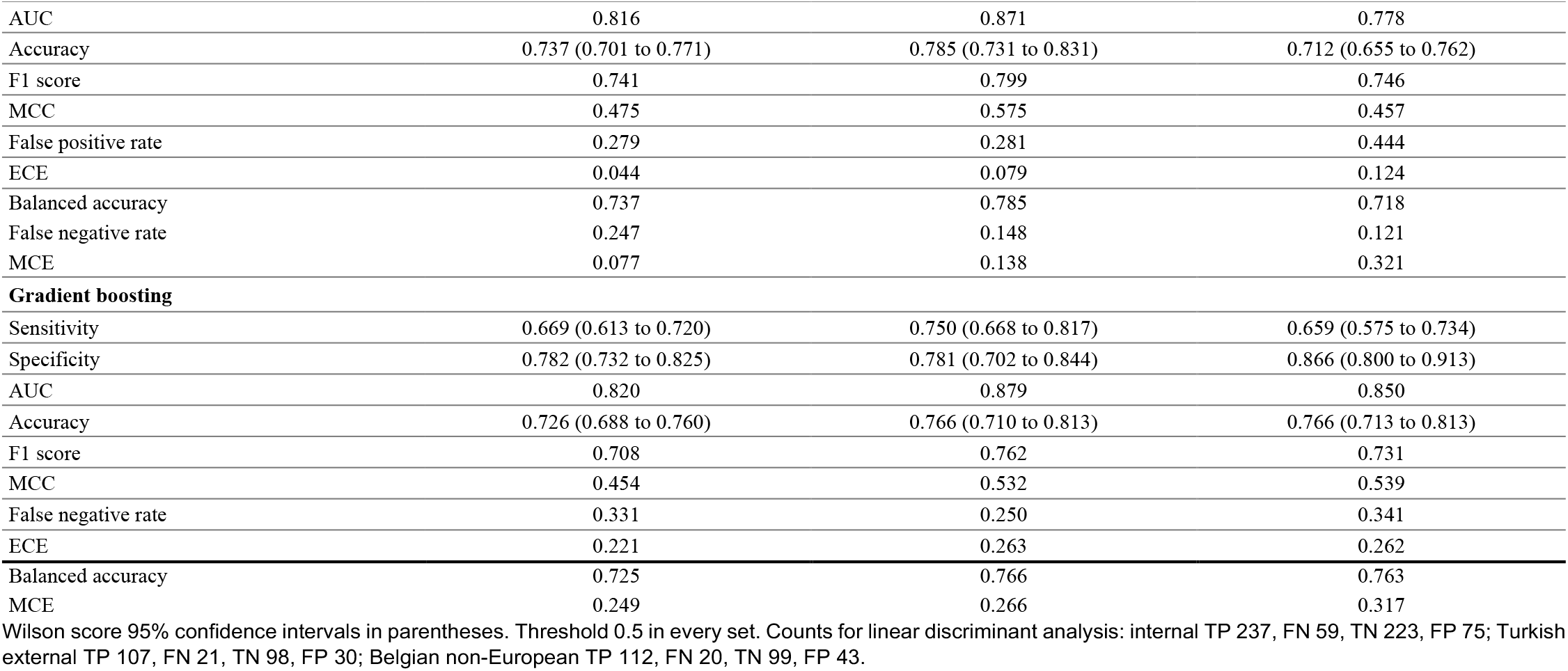
Class iii task, performance of the three retained models.

|  | Internal validation (n = 594) | Turkish external (n = 256) | Belgian non-European (n = 274) |
| --- | --- | --- | --- |
| **Linear discriminant analysis** |  |  |  |
| Sensitivity | 0.801 (0.751 to 0.842) | 0.836 (0.762 to 0.890) | 0.848 (0.778 to 0.900) |
| Specificity | 0.748 (0.696 to 0.794) | 0.766 (0.685 to 0.831) | 0.697 (0.617 to 0.767) |
| AUC | 0.845 | 0.902 | 0.835 |
| Accuracy | 0.774 (0.739 to 0.806) | 0.801 (0.748 to 0.845) | 0.770 (0.717 to 0.816) |
| Balanced accuracy | 0.774 | 0.801 | 0.773 |
| Precision | 0.760 (0.709 to 0.804) | 0.781 (0.705 to 0.842) | 0.723 (0.647 to 0.787) |
| F1 score | 0.780 | 0.808 | 0.780 |
| MCC | 0.550 | 0.603 | 0.550 |
| False positive rate | 0.252 | 0.234 | 0.303 |
| False negative rate | 0.199 | 0.164 | 0.152 |
| ECE | 0.039 | 0.053 | 0.091 |
| MCE | 0.133 | 0.148 | 0.205 |
| **Logistic regression** |  |  |  |
| Sensitivity | 0.753 (0.701 to 0.799) | 0.852 (0.780 to 0.903) | 0.879 (0.812 to 0.924) |
| Specificity | 0.721 (0.668 to 0.769) | 0.719 (0.635 to 0.789) | 0.556 (0.474 to 0.635) |
| AUC | 0.816 | 0.871 | 0.778 |
| Accuracy | 0.737 (0.701 to 0.771) | 0.785 (0.731 to 0.831) | 0.712 (0.655 to 0.762) |
| F1 score | 0.741 | 0.799 | 0.746 |
| MCC | 0.475 | 0.575 | 0.457 |
| False positive rate | 0.279 | 0.281 | 0.444 |
| ECE | 0.044 | 0.079 | 0.124 |
| Balanced accuracy | 0.737 | 0.785 | 0.718 |
| False negative rate | 0.247 | 0.148 | 0.121 |
| MCE | 0.077 | 0.138 | 0.321 |
| **Gradient boosting** |  |  |  |
| Sensitivity | 0.669 (0.613 to 0.720) | 0.750 (0.668 to 0.817) | 0.659 (0.575 to 0.734) |
| Specificity | 0.782 (0.732 to 0.825) | 0.781 (0.702 to 0.844) | 0.866 (0.800 to 0.913) |
| AUC | 0.820 | 0.879 | 0.850 |
| Accuracy | 0.726 (0.688 to 0.760) | 0.766 (0.710 to 0.813) | 0.766 (0.713 to 0.813) |
| F1 score | 0.708 | 0.762 | 0.731 |
| MCC | 0.454 | 0.532 | 0.539 |
| False negative rate | 0.331 | 0.250 | 0.341 |
| ECE | 0.221 | 0.263 | 0.262 |
| Balanced accuracy | 0.725 | 0.766 | 0.763 |
| MCE | 0.249 | 0.266 | 0.317 |

Wilson score 95% confidence intervals in parentheses. Threshold 0.5 in every set. Counts for linear discriminant analysis: internal TP 237, FN 59, TN 223, FP 75; Turkish external TP 107, FN 21, TN 98, FP 30; Belgian non-European TP 112, FN 20, TN 99, FP 43.

Specificity was 0.748 on internal validation, 0.766 in the Turkish external cohort and 0.697 in the Belgian non-European external cohort. The difference between the two external cohorts was 0.068 (95% CI −0.038 to 0.171; adjusted p = 0.618). The false positive rate was 0.234 and 0.303 respectively.

AUC was 0.902 in the Turkish external cohort and 0.835 in the Belgian non-European external cohort (Fig. 1). The difference between the two external cohorts was 0.066 (95% CI 0.006 to 0.128; p = 0.029). The magnitude of the difference between the two external cohorts varied across measures, from 0.027 for the F1 score to 0.068 for specificity. Between cohort differences in the primary outcome measures are given in Table IV. Maximum calibration error was higher in the Belgian non-European cohort for all three models in the Class III task; rising from 0.133 on internal validation to 0.205 for linear discriminant analysis and from 0.077 to 0.321 for logistic regression.

**TABLE IV.** Between cohort differences in primary outcome measures.

| Comparison | Difference (95% CI) | z | Adjusted p |
| --- | --- | --- | --- |
| **Class III task, specificity** |  |  |  |
| Internal vs Turkish external | -0.017 (-0.101 to 0.075) | -0.38 | 1.000 |
| Internal vs Belgian non-Eur. | 0.051 (-0.036 to 0.143) | 1.13 | 0.773 |
| Turkish ext. vs Belgian non-Eur. | 0.068 (-0.038 to 0.171) | 1.26 | 0.618 |
| **Class III task, sensitivity** |  |  |  |
| Internal vs Turkish external | -0.035 (-0.108 to 0.049) | -0.85 | 1.000 |
| Internal vs Belgian non-Eur. | -0.048 (-0.119 to 0.034) | -1.18 | 0.717 |
| Turkish ext. vs Belgian non-Eur. | -0.013 (-0.102 to 0.077) | -0.28 | 1.000 |
| **Class III task, AUC** |  |  |  |
| Turkish ext. vs Belgian non-Eur. | 0.066 (0.006 to 0.128) |  | 0.029 |
| **Class I and II task, specificity** |  |  |  |
| Internal vs Turkish external | -0.207 (-0.278 to -0.090) | -3.17 | 0.005 |
| Internal vs Belgian non-Eur. | -0.122 (-0.178 to -0.061) | -3.88 | <0.001 |
| Turkish ext. vs Belgian non-Eur. | 0.085 (-0.033 to 0.161) | 1.49 | 0.407 |
| **Class I and II task, sensitivity** |  |  |  |
| Internal vs Turkish external | -0.034 (-0.118 to 0.058) | -0.74 | 1.000 |
| Internal vs Belgian non-Eur. | 0.087 (0.023 to 0.150) | 2.68 | 0.022 |
| Turkish ext. vs Belgian non-Eur. | 0.121 (0.024 to 0.210) | 2.45 | 0.043 |
| **Class I and II task, AUC** |  |  |  |
| Turkish ext. vs Belgian non-Eur. | 0.152 (0.089 to 0.211) |  | <0.001 |

Proportions: Newcombe hybrid score intervals, two proportion z tests, Bonferroni corrected for three comparisons per measure. AUC: bootstrap percentile intervals,

**Fig. 1.**
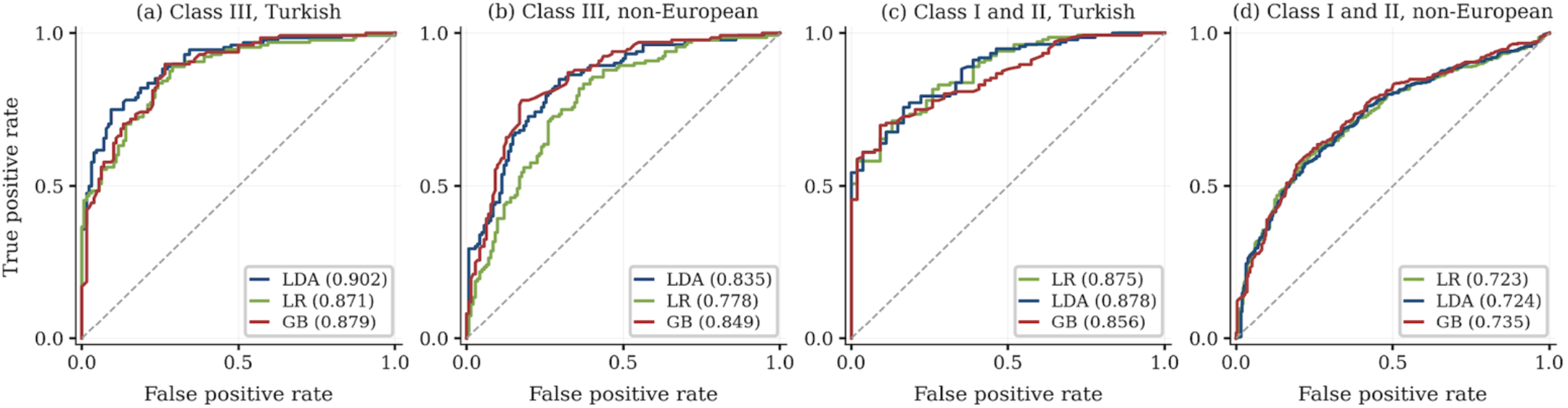
Receiver operating characteristic curves for the three retained models in each task and external cohort. (a) Class III task, Turkish external cohort. (b) Class III task, Belgian non-European cohort. (c) Class I and Class II task, Turkish external cohort. (d) Class I and Class II task, Belgian non-European cohort. LDA, linear discriminant analysis; LR, logistic regression; GB, gradient boosting. Values in parentheses are the AUC.

A subgroup analysis by age was conducted for the primary model in the Class III task. In the Turkish external cohort, adults (n = 148; 86 Class III, 62 non-Class III) showed higher sensitivity than children (n = 108; 42 Class III, 66 non-Class III): 0.89 versus 0.71, with corresponding false negative rates of 0.11 and 0.29. F1 scores were 0.83 for adults and 0.76 for children. In the Belgian non-European cohort, adults (n = 131; 61 Class III, 70 non-Class III) similarly showed higher sensitivity than children (n = 151; 71 Class III, 80 non-Class III): 0.90 versus 0.80, with false negative rates of 0.10 and 0.20. F1 was 0.79 for adults and 0.76 for children. Age-stratified analyses were performed on independently age-balanced subsamples (Turkish external: n = 256; Belgian non-European: n = 282) that differ from the main class-balanced cohorts used elsewhere; the Belgian subsample comprised 132 Class III and 150 non-Class III cases. Across both cohorts, children showed a false negative rate approximately 0.15 to 0.18 higher than adults. F1 was identical in children across the two cohorts (0.76 in both), while adults showed a small cross-cohort decline from 0.83 to 0.79. Full age-stratified metrics are given in Table V.

**TABLE V.** Age-Stratified Class III Task Performance of the Primary Model (LDA)

| Metric | Turkish external cohort |  | Belgian non-European cohort |  |
| --- | --- | --- | --- | --- |
|  | Adults (n = 148) | Children (n = 108) | Adults (n = 131) | Children (n = 151) |
| **Class III (positive)** | 86 | 42 | 61 | 71 |
| Non-Class III (negative) | 62 | 66 | 70 | 80 |
| **Sensitivity** | **0.89** | 0.71 | **0.90** | 0.80 |
| False negative rate | 0.11 | 0.29 | 0.10 | 0.20 |
| False positive rate | 0.27 | 0.17 | 0.33 | 0.27 |
| **F1 score** | **0.83** | 0.76 | 0.79 | 0.76 |
| ECE | 0.07 | 0.07 | 0.16 | 0.06 |
| MCE | 0.24 | 0.34 | 0.37 | 0.13 |

Adults: age $\geq 18$ yr; Children: age $< 18$ yr. F1 was identical in children across both cohorts (0.76 in both). Age-stratified subsamples were independently balanced for adult/child composition within each class; the Belgian non-European age-stratified cohort (n = 282: 132 Class III, 150 non-Class III) differs from the main class-balanced cohort (n = 274: 132 Class III, 142 non-Class III) used in Tables III, IV and VII.

Class I and Class II task results are given in Table VI. Sensitivity was 0.628 on internal validation, 0.662 in the Turkish external cohort and 0.541 in the Belgian non-European external cohort. Specificity was 0.682, 0.889 and 0.804, respectively. The difference in specificity between the two external cohorts was 0.085 (95% CI −0.033 to 0.161; adjusted p = 0.407). The Turkish external cohort contained 54 Class I cases. AUC was 0.706 on internal validation, 0.875 in the Turkish external cohort and 0.723 in the Belgian non-European external cohort, a difference between the external cohorts of 0.152 (95% CI 0.089 to 0.211; p < 0.001). The F1 score differed by 0.139 between the two external cohorts.

**TABLE VI.**
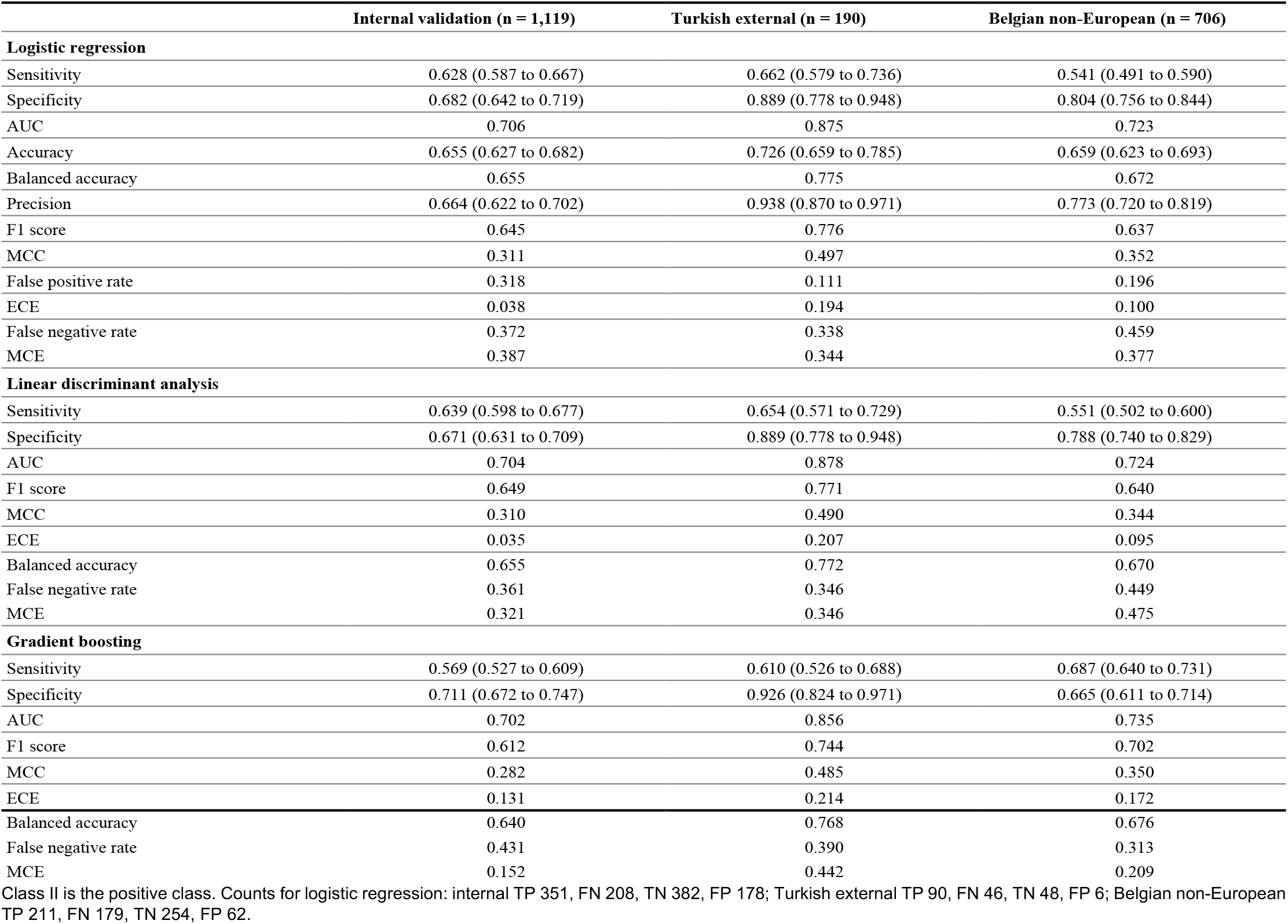
Class I AND Class ii task, performance of the three retained models.

|  | Internal validation (n = 1,119) | Turkish external (n = 190) | Belgian non-European (n = 706) |
| --- | --- | --- | --- |
| **Logistic regression** |  |  |  |
| Sensitivity | 0.628 (0.587 to 0.667) | 0.662 (0.579 to 0.736) | 0.541 (0.491 to 0.590) |
| Specificity | 0.682 (0.642 to 0.719) | 0.889 (0.778 to 0.948) | 0.804 (0.756 to 0.844) |
| AUC | 0.706 | 0.875 | 0.723 |
| Accuracy | 0.655 (0.627 to 0.682) | 0.726 (0.659 to 0.785) | 0.659 (0.623 to 0.693) |
| Balanced accuracy | 0.655 | 0.775 | 0.672 |
| Precision | 0.664 (0.622 to 0.702) | 0.938 (0.870 to 0.971) | 0.773 (0.720 to 0.819) |
| F1 score | 0.645 | 0.776 | 0.637 |
| MCC | 0.311 | 0.497 | 0.352 |
| False positive rate | 0.318 | 0.111 | 0.196 |
| ECE | 0.038 | 0.194 | 0.100 |
| False negative rate | 0.372 | 0.338 | 0.459 |
| MCE | 0.387 | 0.344 | 0.377 |
| **Linear discriminant analysis** |  |  |  |
| Sensitivity | 0.639 (0.598 to 0.677) | 0.654 (0.571 to 0.729) | 0.551 (0.502 to 0.600) |
| Specificity | 0.671 (0.631 to 0.709) | 0.889 (0.778 to 0.948) | 0.788 (0.740 to 0.829) |
| AUC | 0.704 | 0.878 | 0.724 |
| F1 score | 0.649 | 0.771 | 0.640 |
| MCC | 0.310 | 0.490 | 0.344 |
| ECE | 0.035 | 0.207 | 0.095 |
| Balanced accuracy | 0.655 | 0.772 | 0.670 |
| False negative rate | 0.361 | 0.346 | 0.449 |
| MCE | 0.321 | 0.346 | 0.475 |
| **Gradient boosting** |  |  |  |
| Sensitivity | 0.569 (0.527 to 0.609) | 0.610 (0.526 to 0.688) | 0.687 (0.640 to 0.731) |
| Specificity | 0.711 (0.672 to 0.747) | 0.926 (0.824 to 0.971) | 0.665 (0.611 to 0.714) |
| AUC | 0.702 | 0.856 | 0.735 |
| F1 score | 0.612 | 0.744 | 0.702 |
| MCC | 0.282 | 0.485 | 0.350 |
| ECE | 0.131 | 0.214 | 0.172 |
| Balanced accuracy | 0.640 | 0.768 | 0.676 |
| False negative rate | 0.431 | 0.390 | 0.313 |
| MCE | 0.152 | 0.442 | 0.209 |

Class II is the positive class. Counts for logistic regression: internal TP 351, FN 208, TN 382, FP 178; Turkish external TP 90, FN 46, TN 48, FP 6; Belgian non-European TP 211, FN 179, TN 254, FP 62.

AUC was lower in the Belgian non-European external cohort than in the Turkish external cohort for all three retained models in both tasks. The difference reached significance in five of the six comparisons (Table VII).

**TABLE VII.** AUC in the two external cohorts across models.

| Task | Model | Turkish ext. | Belgian non-Eur. | Difference (95% CI) | p |
| --- | --- | --- | --- | --- | --- |
| Class III | Linear discriminant analysis | 0.902 | 0.835 | 0.066 (0.006 to 0.128) | 0.029 |
| Class III | Logistic regression | 0.871 | 0.778 | 0.093 (0.024 to 0.163) | 0.009 |
| Class III | Gradient boosting | 0.879 | 0.850 | 0.030 (-0.030 to 0.090) | 0.337 |
| Class I and II | Logistic regression | 0.875 | 0.723 | 0.152 (0.089 to 0.211) | <0.001 |
| Class I and II | Linear discriminant analysis | 0.878 | 0.724 | 0.154 (0.090 to 0.213) | <0.001 |
| Class I and II | Gradient boosting | 0.856 | 0.735 | 0.122 (0.056 to 0.182) | <0.001 |

Bootstrap percentile intervals with 10,000 replicates. AUC was lower in the Belgian non-European external cohort in all six comparisons.

The Turkish external cohort was also evaluated without class balancing (n = 318, 40.3% Class III). Sensitivity was unchanged at 0.836, since the Class III group was identical in both versions. Specificity was 0.837 and AUC 0.924, giving differences from the Belgian non-European external cohort of 0.140 (95% CI 0.048 to 0.232; adjusted p = 0.008) and 0.088 (95% CI 0.034 to 0.145; p = 0.002) respectively.

## IV. Discussion

Models developed on Turkish photographic data were applied to two external cohorts using the same saved model files and the same decision threshold. Performance was retained in the Turkish external cohort. In the Belgian non-European external cohort, discrimination was lower, and in the Class III task specificity was lower while sensitivity was unchanged. Lower discrimination was observed in both tasks and in all three models tested. In a screening application, failing to detect an affected patient and flagging an unaffected one are not clinically equivalent. A child with a developing skeletal Class III who is not flagged may pass the growth period in which orthopedic correction is most effective [7], [8], whereas a child flagged in error undergoes a specialist examination that proves unnecessary.

The feature extraction and classification approach was selected empirically. A convolutional neural network with a pretrained backbone underperformed geometric feature classifiers on this development set, a result consistent with comparisons of classical and deep learning classifiers at similar data scales [17], [18]. Encoding the relationship between facial landmarks and skeletal anatomy as explicit geometric measurements, rather than asking a model to recover it from pixel level data, appears to be the more data efficient approach at current training set sizes. Whether deep learning would match this approach with larger training corpora is a question for future work.

Because the Belgian non-European external analysis set was matched to the development set in prevalence and age composition, differences in case mix appear unlikely to account for the difference observed between the two external cohorts.

Both comparison cohorts were external, in that neither contributed any case to training. The Turkish external cohort was also drawn from a different institution, yet performance there was maintained. Institutional difference alone therefore does not account for the result observed in the Belgian non-European external cohort.

The two tasks used different targets, different selected features and different top ranked models. Three models were tested within each task. All six comparisons showed lower discrimination in the Belgian non-European external cohort. A difference in image acquisition or feature extraction specific to one cohort would be expected to affect both tasks. A property specific to one task or one model would not appear six times.

In the Class I and Class II task, both external cohorts showed higher discrimination than internal validation. This illustrates why the comparison between the two external cohorts, rather than between internal and external performance, is the informative one.

The secondary measures behaved differently from the primary ones. In the Class III task the F1 score differed by 0.027 between the two external cohorts, while AUC differed by 0.066 over the same comparison. The F1 score combines sensitivity and precision and does not include specificity. Where sensitivity is preserved and specificity declines, it will therefore remain close to constant.

The model continued to identify affected patients in the Class III task, and sensitivity was marginally higher in the Belgian non-European external cohort, while classification of unaffected patients declined. The model assigned a Class III label to a greater proportion of patients who did not have the condition. Because the model estimates a skeletal outcome from facial morphology, its behavior depends on the relationship between the two, and this relationship varies between populations [13], [14]. The morphological basis of the difference observed here was not measured in the present study.

A recent review of artificial intelligence in orthodontics reported that models developed in single centers lost up to 20% of their accuracy when tested across institutions and identified the limited diversity of development datasets and the absence of independent external validation as recurring problems across task domains [15]. The present study addresses both. Similar losses on external application have been documented in other imaging domains including prostate MRI [24], with individual reports of considerably greater degradation [19]. Noseworthy et al. reported that a deep learning model performed well in one population group while showing reduced performance in others and attributed this to limited diversity in the development data [16].

A pre-specified sensitivity analysis evaluated whether the observed performance pattern could be explained by the difference in class prevalence between training and external validation rather than by a genuine population-specific effect. The Turkish external cohort was evaluated a second time without class balancing (n = 318, 40.3% Class III). Sensitivity was unchanged at 0.836, since the Class III group was identical in both evaluations. Specificity rose from 0.766 to 0.837 as expected from the larger pool of true-negative cases, and AUC rose from 0.902 to 0.924. The signature of prior-probability shift was absent. Sensitivity was preserved and the false-positive rate was not inflated. Sensitivity was marginally higher in the Belgian non-European cohort (0.848) than in either Turkish external cohort evaluation, confirming that the model retained its ability to detect affected patients across populations and that the observed specificity reduction was a property of the population rather than an artefact of class balancing.

The three development sites contributed cases with similar Class III prevalence (51.5%, 51.2% and 44.8%), and the Turkish external cohort was drawn from an institution that contributed nothing to training. Cases were not labelled by site after pooling, so a breakdown of model performance by development site was not estimable. Similar class prevalence across sites does not exclude other sources of site-level variation, differences in image acquisition conditions, patient demographic composition beyond class prevalence, or inter-rater labelling conventions, and these cannot be ruled out as partial contributors to the pattern observed in the Belgian non-European cohort.

The ANB angle was used as the reference standard in every cohort. It is the most widely used cephalometric indicator of the sagittal skeletal relationship, and in a comparison of six analyses it showed the highest agreement with the consensus diagnosis and the highest positive predictive value in the Class II and Class III groups [25]. The thresholds applied to it, however, derive from norms established in predominantly Caucasian samples. Cephalometric norms differ between populations: African American adults show a larger ANB than published Caucasian values [26], and in African Brazilian adults with excellent occlusion, seven angular and six linear measurements differed significantly from European-American standards, leading the authors to conclude that Caucasian norms should not be applied to that population [27]. Turkish adults and adolescents likewise differ from their North American counterparts in facial morphology [13], [14].

A model trained to reproduce a Caucasian-derived threshold in a Turkish population, and then applied to a non-European one, is therefore evaluated against a criterion that may not partition the two populations in the same way. Part of what we have measured as reduced specificity may reflect the population dependence of the reference standard rather than of the model alone. The two are not separable in the present design, since establishing which of them is responsible would require population-specific skeletal criteria, or a criterion independent of population norms, applied to the same patients. This does not alter the practical conclusion. Whether the source is the model, the criterion, or both, a screening tool that carries a Caucasian-derived threshold into a population for which that threshold was not established will behave differently there, and that behavior has to be measured rather than assumed.

Maximum calibration error diverged from expected calibration error in the Belgian non-European cohort. Logistic regression showed an MCE of 0.321 against an ECE of 0.124, and the pattern was consistent across models. A large difference between MCE and ECE indicates that miscalibration is not distributed evenly across the probability scale but is concentrated in specific ranges. Where a tool’s output is used to guide referral decisions, a patient assigned a score of 0.7 expects the model to be right about 70% of the time, so that a failure of calibration at that point makes the resulting decision less reliable than the score implies. This finding adds a reason, beyond discrimination alone, to recommend population-specific evaluation of calibration before deployment.

The observed pattern is here described as a finding about cross-population generalizability rather than algorithmic bias. Algorithmic bias, in the strict sense, requires evidence that a model produces systematically different outcomes for identifiable subgroups relative to a ground truth that treats those subgroups equivalently. In the present study the ground truth itself, an ANB threshold derived from Caucasian normative data, is not demonstrably equivalent across populations, and the observed specificity reduction is consistent with a difference in the relationship between facial morphology and the cephalometric criterion rather than with a differential error by the model. Sensitivity was preserved in the Class III task, and lower discrimination recurred in both tasks and across all three models. These observations are consistent with a generalizability finding and do not establish bias in the strict sense.

The subgroup analysis by age revealed a consistent pattern across both external cohorts, with sensitivity substantially lower in children than in adults and false negative rates of 0.29 and 0.20 in children against 0.11 and 0.10 in adults in the Turkish external and Belgian non-European cohorts respectively. This is clinically significant because the primary target of early orthodontic screening is the pediatric population, yet the model performed markedly less well in children than in adults across both cohorts and independent of population of origin. One candidate explanation is that the skeletal discrepancy itself is less pronounced in children. In untreated Class III subjects, the mandible continues to outgrow the maxilla throughout the growth period, ANB becomes progressively more negative from year to year, and a substantial part of this differential growth occurs after the adolescent growth spurt [28]. A child who will meet the Class III criterion as an adult may therefore lie close to the threshold at the age at which screening takes place, with correspondingly limited expression in the facial profile, so that geometric feature values overlap more between classes at younger ages. Another is that the development set contained 60% adults, which may have biased the model toward adult facial proportions. The identical F1 in children across the two external cohorts (0.76 in both) suggests that the cross-population difference in aggregate performance reflects the higher proportion of children in the Belgian non-European cohort rather than a population-specific failure in children. Future development should consider age-stratified model training or a pediatric-specific classifier.

Generalizability is a property of a model in relation to a target population. The same models performed as expected within their development population and less well outside it. An accuracy figure reported without the population in which it was obtained gives limited information about performance elsewhere.

## V. Limitations

Age composition differed between cohorts, with adults comprising 67% of Class III cases in the Turkish external cohort and 46% in the Belgian non-European cohort. A subgroup analysis showed that sensitivity was lower in children than in adults across both cohorts (0.71–0.80 versus 0.89–0.90), and the higher proportion of children in the Belgian non-European cohort (54% versus 33%) contributes to its lower aggregate performance. The between-cohort difference in F1 was absent in children (0.76 in both cohorts) and small in adults (0.83 versus 0.79), suggesting that age composition accounts for part of the aggregate cross-cohort difference.

Individual age values were unavailable for the analysis sets, and age composition is reported as the proportion of adults rather than as a mean. Sex was not recorded in the anonymized datasets.

The Belgian non-European external cohort was recruited at a single clinic in Brussels. It comprised 873 patients identified as being non-European by an orthodontist on the basis of facial appearance, rather than by self-report, genetic testing or random selection. Selection effects therefore cannot be excluded, although sensitivity did not differ between cohorts in the Class III task and lower discrimination recurred in a second task. The findings cannot be extended to non-Turkish or non-European populations in general based on one cohort at one site.

Site of origin is recorded for the development pools but not for the individual cases retained after balancing, so a within-population, between-site breakdown of model performance was not possible. Class III prevalence across the three contributing sites was 51.5%, 51.2% and 44.8%.

In the Class I and Class II task the Belgian non-European external cohort was neither prevalence balanced nor age matched, and the two tasks differ in this respect. Maximum calibration error in the Class I and Class II task was high across all models and cohorts, including internal validation, suggesting that probability outputs of the Class I and Class II classifier should be used with caution regardless of population. The binary label may be more reliable than the associated probability in this task.

## VI. Conclusion

Machine learning models developed on Turkish photographic data retained their performance in an independent Turkish cohort and showed lower discrimination in the Belgian non-European external cohort, with lower specificity and unchanged sensitivity in the Class III task. Lower discrimination was reproduced in both tasks and in all three models tested in each. The F1 score differed little between the two external cohorts, and the difference would not have been apparent had that measure been used alone.

The model was consistent within its development population and less outside it. This is not a property of one system. Photograph-based tools are now in use without published information on the populations from which their development data were drawn, and a user has no means of knowing whether the population they belong to was represented. Where such a tool assigns a label from a facial photograph, the composition of the data behind that label determines who it will serve accurately and who it will not. Validation in the population where a tool is to be deployed should therefore be part of its development rather than a step that follows it. Calibration varied between cohorts and was poorest in the Belgian non-European external cohort; maximum calibration error reached 0.321 for logistic regression, indicating that miscalibration was concentrated in specific probability ranges. Population-specific calibration assessment should accompany discrimination metrics before clinical deployment. Future work should extend this design to further populations, record self-reported ethnicity, age, and sex at the point of collection so that subgroup analysis becomes possible, and report discrimination and calibration alongside prevalence independent measures.

## Data Availability

The data analysed in this study consist of pre-treatment facial photographs and lateral cephalograms of orthodontic patients. Because facial images are potentially identifying, the raw data cannot be made publicly available or deposited in a repository. Anonymized derived data may be made available on reasonable request to the corresponding author, subject to approval by the Bezmialem Vakif University research ethics committee and by the institutions that provided the data.

## Acknowledgment

Belgian clinical data were collected at PEBS Orthodontics (Brussels, Belgium). The funding sources and PEBS Orthodontics had no role in study design, analysis, interpretation, or the decision to submit for publication.

Generative artificial intelligence tools were used during the preparation of this manuscript for language editing, clarity improvement of the written text and to format the tables. All scientific content, data analysis, results, interpretation, and conclusions are the sole responsibility of the authors. No AI tool was used to generate data, figures, statistical analyses, or any part of the Results or Discussion sections.

## Conflicts of interest

Banu Kiliç and Tuğba Önal-Süzek are co-founders and co-owners of Kedi Mobil Uygulama Anonim Şirketi (https://kedi-mobile.com), the company that developed and operates the screening application evaluated in this study, and receive salary support funded through the TÜBİTAK 1507 SME R&D Support Programme (grant no. 7248072) and the European Union’s Horizon Europe Women TechEU initiative (grant agreement no. 101113949). Mehmet Cihan Sakman and Özgür Kiliç receive salary support from Kedi Mobil Uygulama Anonim Şirketi. The machine learning models evaluated in this study are deployed in a free mobile application and a free web-based screening interface operated by the company; neither generates revenue from the screening function. Emmanuel Jr Dumu is employed by PEBS Orthodontics, Brussels, Belgium, which provided the clinical setting and patient data for the Belgian non-European external cohort. PEBS Orthodontics had no role in study design, data analysis, interpretation, or the decision to publish. Emmanuel Jr Dumu contributed as an independent clinician responsible for patient data collection. The remaining authors declare no conflicts of interest.

## Notes

This work was supported by the TÜBİTAK 1507 SME R&D Support Programme under Grant 7248072 and by the European Union’s Horizon Europe research and innovation programme under the Women TechEU initiative under Grant Agreement 101113949.

### Competing Interest Statement

Conflict of Interest Statement
Photograph-Based Skeletal Malocclusion Screening with Geometric Machine Learning: External Validation Across Turkish and Non-European Cohorts
Banu Kilic, Tugba Onal Suzek, Mehmet Cihan Sakman, and Ozgur Kilic received salary support from Kedi Mobil Uygulama Anonim Sirketi (https://kedi-mobile.com), fully funded through TUBİTAK 1507 SME R&D Support Programme grant no. 7248072. Banu Kilic and Tugba Onal-Suzek additionally hold equity in the company. The machine learning models evaluated in this study are deployed in a free mobile application and a free web-based screening interface operated by the company; neither generates revenue from the screening function. Emmanuel Jr Dumu is employed by PEBS Orthodontics, Brussels, Belgium, which provided the clinical setting and patient data for the Belgian non-European external cohort. PEBS Orthodontics had no role in study design, interpretation, or the decision to publish. Emmanuel Jr Dumu contributed as an independent clinician responsible for patient data collection. The remaining authors declare no conflicts of interest.

### Clinical Trial

NCT07162753

### Clinical Protocols

https://clinicaltrials.gov/study/NCT07162753

### Author Declarations

The Non-Interventional Research Ethics Committee of Bezmialem Vakif University, Istanbul, Turkey gave ethical approval for this work (decision numbers 2024/342 on 04 September 2024 and 2024/438 on 11 December 2024).

